# Prediction of severe COVID-19 infection at the time of testing: A machine learning approach

**DOI:** 10.1101/2021.10.15.21264970

**Authors:** Faraz Khoshbakhtian, Ardian Lagman, Dionne M. Aleman, Randy Giffen, Proton Rahman

**Affiliations:** Department of Mechanical & Industrial Engineering, University of Toronto, Toronto, Ontario, Canada; IBM Canada, Toronto, ON, Canada; Eastern Health, Newfoundland & Labrador, Canada

## Abstract

Early and effective detection of severe infection cases during a pandemic can significantly help patient prognosis and resource allocation. We develop a machine learning framework for detecting severe COVID-19 cases at the time of RT-PCR testing. We retrospectively studied 988 patients from a small Canadian province that tested positive for SARS-CoV-2 where 42 (4%) cases were *at-risk* (i.e., resulted in hospitalization, admission to ICU, or death), and 8 (< 1%) cases resulted in death. The limited information available at the time of RT-PCR testing included age, comorbidities, and patients’ reported symptoms, totaling 27 features. Vaccination status was unavailable. Due to the severe class imbalance and small dataset size, we formulated the problem of detecting severe COVID as anomaly detection and applied three models: one-class support vector machine (OCSVM), weight-adjusted XGBoost, and weight-adjusted AdaBoost. The OCSVM was the best performing model for detecting the deceased cases with an average 95% true positive rate (TPR) and 27.2% false positive rate (FPR). Meanwhile, the XGBoost provided the best performance for detecting the at-risk cases with an average 96.2% TPR and 19% FPR. In addition, we developed a novel extension to SHAP interpretability to explain the outputs from the models. In agreement with conventional knowledge, we found that comorbidities were influential in predicting severity, however, we also found that symptoms were generally more influential, noting that machine learning combines all available data and is not a single-variate statistical analysis.

## 1 Introduction

The COVID pandemic showed that even the most resilient national healthcare systems are prone to quickly becoming overwhelmed in times of widespread infectious disease outbreaks, especially when there is no known effective treatment for the disease [9, 16, 22]. Early detection of severe COVID cases, that is, those resulting in hospitalization, ICU, or death, can help plan and manage scarce resources to best treat patients and manage scarce health care resources. We developed machine learning tools that can detect severe COVID cases at the time of RT-PCR testing, before patients present at emergency departments, are seen by any clinical practitioners, or have any diagnostic analysis performed. Outcome prediction at this earliest possible moment of knowledge of COVID infection is unique among the literature predicting COVID severity, which all make predictions after a patient has already presented at an emergency department. Additionally, our focus on a small regional area may help avoid underlying biases in the dataset and ensure the predictive models are tuned for the actual patient population being treated.

Many studies have shown the promise of machine learning in augmenting decision-making capabilities for clinicians in diagnosing COVID through imaging data [17, 25, 30] and tabular data [1, 29]. The use cases range from detecting COVID positive cases to detecting severe cases from the infected population. To the best of our knowledge, machine learning applications for COVID prognosis have exclusively focused on supervised classification methods. Even though the reported metrics show high efficacy, they generally require somewhat large labeled datasets—18000 patients in Subudhi et al. [29] and 6995 in Assaf et al. [1]—and all rely on blood test and/or imaging data not available until a patient has already presented to an emergency department or an out-patient clinic. In the case of a global pandemic, large labeled datasets indicate human loss and missed opportunities to save lives, as well as are usually not available for single centers or individual regions to analyze and make their own predictive models. Further, datasets aggregated over numerous hospitals run the risk of biasing predictions against actual patient populations seen in smaller regional areas.

Model interpretability and decision explainability are essential parts of any health-oriented machine learning framework that aims for clinical relevance. As machine learning models become more complex, it becomes harder to explain and fully understand the reasons behind their decisions. However, especially in medical applications, it is crucial not only to verify the performance of the models but also to make sure the decisions made by them are explainable and derived from relevant pieces of information. On the one hand, biases can quickly propagate from datasets to models and onto the medical institutions. On the other hand, mere outcome predictions cannot help medical practitioners save their patients’ lives. Feature interpretability and output explainability allow for assessing models’ understanding of risk factors, which gives medical practitioners insight to alleviate those risk factors.

Thus, we develop an unsupervised anomaly detection and imbalance-aware supervised avenues for early risk assessment and prognosis of COVID patients, and use interpretability techniques to explain the impact of patient features on the severity predictions. Given the relative rarity of severe COVID outcomes among all COVID-positive patients, we formulate the problem of detecting severe COVID as one of anomaly detection. We train two supervised models—weight-adjusted XGBoost and AdaBoost—and one unsupervised model—a one-class support vector machine (OCSVM)—to detect at-risk cases (hospitalization, ICU, death) as well as deceased cases. Despite the limited data available at the time of RT-PCR testing, which includes age, comorbidities, patient-reported symptoms (but not vaccination status), our models show high sensitivity to the at-risk patients even when trained on a relatively small dataset. We use SHapley Additive exPlanations (SHAP) [21] to interpret model decisions based on understandable and intuitive features and feature groups, allowing for validation by clinicians. We novelly extend SHAP calculations to the particular needs of our dataset and modeling approaches, which include feature groups in addition to individual features (e.g., a “symptoms group” feature in addition to individual specific symptoms) and aggregate SHAP analysis across multiple cross-fold validations, as needed by our small dataset. Our framework can be used in the early and later stages of regional outbreaks to closely monitor the at-risk patients and plan for resource allocation.

## 2 Literature review

Since the start of the COVID pandemic in early 2020, a myriad of works have shown that machine learning can be an essential and effective tool in alleviating the risks and costs of a global pandemic. Both structured and unstructured data can be used to diagnose patients and/or perform prognostic assessments at different stages of their infection. These frameworks can help inform decision-making for individual COVID patients at the micro-level or integrate into the macro-level planning and early alert infrastructure.

Some studies take advantage of unstructured data such as radiographic images. These studies mostly use either chest X-rays (CXR) [17, 25, 30] or CT scans [20]; however, CXR images remain a more viable option because of their low cost and widespread adoption. Wang et al. [30] proposed COVID-net, a custom convolutional neural network (CNN) architecture for the classification of COVID infections based on CXR radiography images and report 98.9% positive predictive value (PPV) and 91% sensitivity. CNNs’ performance on image data is almost unchallenged with respect to other machine learning techniques [14, 15, 31]. That said, CNNs are black boxes, and even with advancements in model interpretability, it is still hard for humans to fully understand the decision process of these models [26]. This problem is exacerbated by the need for task-specific and clinically oriented interpretability solutions and a lack of regulations in AI research and development. Therefore, interpretability and decision explainability remain open challenges.

Another body of related machine learning literature uses structured (i.e., tabular) data to assist with pandemic planning and decision making. Brinati et al. [3] showed the feasibility of using machine learning with routine blood test data to achieve similar performance metrics as the gold standard of RT-PCR tests in discriminating between negative and positive COVID cases. Machine learning can also assist in forecasting the severity of the infection for those patients already diagnosed with COVID [12, 18, 29, 32]. Machine learning for prognosis can be used at different stages of COVID infection for patients. Gao et al. [12] used blood test data, comorbidity history, and other basic patient information to assess mortality risk using a voting ensemble of supervised classifiers and achieved high AUC (area under the receiver operating characteristic curve) scores (*>* 90%) across several patient cohorts. Assaf et al. [1] used supervised classification and information at the time of hospital admission to predict the risk of further deterioration. With a small dataset of 162 total hospitalized patients, from which 25 cases deteriorated into critical COVID, they reported a high AUC score of 0.9 that outperformed clinical risk assessment parameters such as APACHE II score [19].

Machine learning classification tasks usually fall under the two categories of supervised and unsupervised learning. Supervised algorithms learn to map the input space onto the output space through encountering labeled data that includes instances of different possible values in the output space [7]. Unsupervised algorithms take on the challenge of learning from unlabeled data and are themselves tasked with discovering reoccurring trends in the data [2]. One the other hand, some machine learning algorithms (that may be supervised or unsupervised) are tasked with classification in severely imbalanced populations; these are called anomaly detection algorithms [4]. We implement AdaBoost [11] and XGBoost [6] for imbalance-aware supervised learning and OCSVM [28] for unsupervised anomaly detection learning. AdaBoost and XGBoost are famous boosting algorithms that bring together many weak classifiers into a single robust classifier [27]. On the other hand, OCSVMs are well-known unsupervised anomaly detection algorithms that can model the decision boundaries of a high-dimensional distribution [8].

Model interpretability for COVID prognosis has been implemented for imaging data, though the interpretability methods used—Grad-CAM in Zhang et al. [33] and Li et al. [20], GSInquire in Wang et al. [30]—provide heatmap overlays that only indicate which parts of an image were important, but not what quality made them important. In the case of structured data, there are numerous model-based and model agnostic methods for interpretability. For example, Brinati et al. [3] trained random forests and leveraged the model-based feature importance. On the other hand, model agnostic interpretability methods are more appropriate when the importances of the same features are being studied across different models.

SHAP is currently one of the most widely adopted interpretability methods; its applications range from COVID prognosis [10, 29] to financial time-series analysis [23]. SHAP draws from game theory to explain the output of any machine learning algorithm by computing the marginal contribution of each data feature in the final output of the model per sample [21]. Notably, SHAP values provide a sample-level understanding of a model’s output by calculating a set of values for each sample and allocating optimal credit to each attribute of that sample. SHAP values are usually calculated only once on the validation samples and are used to study individual feature importances. SHAP does not natively allow for calculations of feature importance across multiple cross-validations, which are often appropriate for small datasets. Additionally, SHAP is designed to assess individual feature importances, however, in our dataset, we create new features that aggregate individual features into groups by clinical interest. Thus, we implemented novel extensions to SHAP for multiple cross-validations and feature groups.

## 3 Data

This retrospective study included all diagnosed SARS-CoV-2 patients from a small Canadian province up until March 26, 2021, under research ethics approval. The diagnosis was confirmed with reverse-transcriptase polymerase chain reaction (RT-PCR) testing. The total number of diagnosed patients in the study was 988, where 42 (4%) cases were at-risk (resulting in hospitalization and/or death), and 8 (< 1%) cases resulted in death. For each patient, information regarding their sex, age, comorbidities, and perceived symptoms at the time of diagnosis were available, totaling 27 features (Table 1). Figure 1 shows the case counts by age, while Figure 2 shows the distribution of comorbidities by age. Distributions of all other features, which include sex, mode of infection acquisition (not included in the modeling attributes), symptoms, and outcome, are shown in Figure 3. We note that the time period of the study spans before and after vaccine availability, however, the data did not contain vaccination status; thus, underlying distributions of features for patients testing positive/negative may have changed during the study, particularly with respect to age.

**Table 1:** Features and feature groups. All features and feature groups other than age are 0/1 values.

| Feature name | Feature groups |  |  |  |  |
| --- | --- | --- | --- | --- | --- |
|  | Comorbid-<br>ities | Constit-<br>utional<br>symptoms | Fever<br>symptoms | Respiratory<br>symptoms | Symptoms<br>combined |
| Age |  |  |  |  |  |
| AMI status | ✓ |  |  |  |  |
| Asthma status | ✓ |  |  |  |  |
| COPD status | ✓ |  |  |  |  |
| Cough |  |  |  | ✓ | ✓ |
| Diarrhea |  |  |  |  | ✓ |
| DM status | ✓ |  |  |  |  |
| Fever |  |  | ✓ | ✓ | ✓ |
| Feverish/Chills |  |  | ✓ | ✓ | ✓ |
| General weakness |  | ✓ |  |  | ✓ |
| Headache |  | ✓ |  |  | ✓ |
| Heart-failure status | ✓ |  |  |  |  |
| Hypertension status | ✓ |  |  |  |  |
| IHD status | ✓ |  |  |  |  |
| Irritability |  | ✓ |  |  | ✓ |
| Loss of appetite |  | ✓ |  |  | ✓ |
| Loss of smell/taste |  |  |  |  | ✓ |
| Nausea |  | ✓ |  |  | ✓ |
| Pain |  | ✓ |  | ✓ | ✓ |
| Painful swallowing |  |  |  |  | ✓ |
| Runny nose |  |  |  |  | ✓ |
| Sex |  |  |  |  |  |
| Shortness of breath |  |  |  | ✓ | ✓ |
| Small red purple spots |  |  |  |  | ✓ |
| Sore throat |  |  |  |  | ✓ |
| Stroke status | ✓ |  |  |  |  |
| Symptomatic |  |  |  |  | ✓ |

AMI: acute myocardial infraction; COPD: chronic obstructive pulmonary disorder; DM: diabetes mellitus; IHD: ischemic heart disease

**Figure 1.**
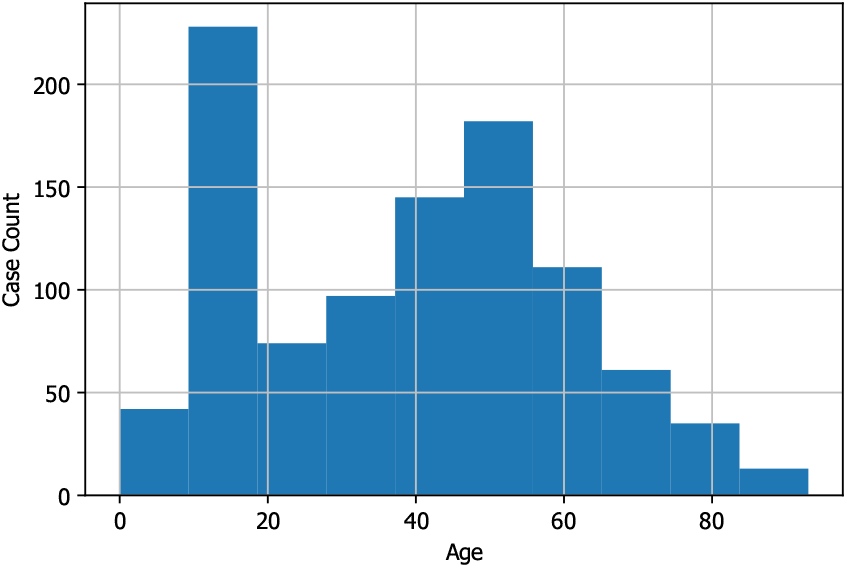
Distribution of patient ages

**Figure 2.**
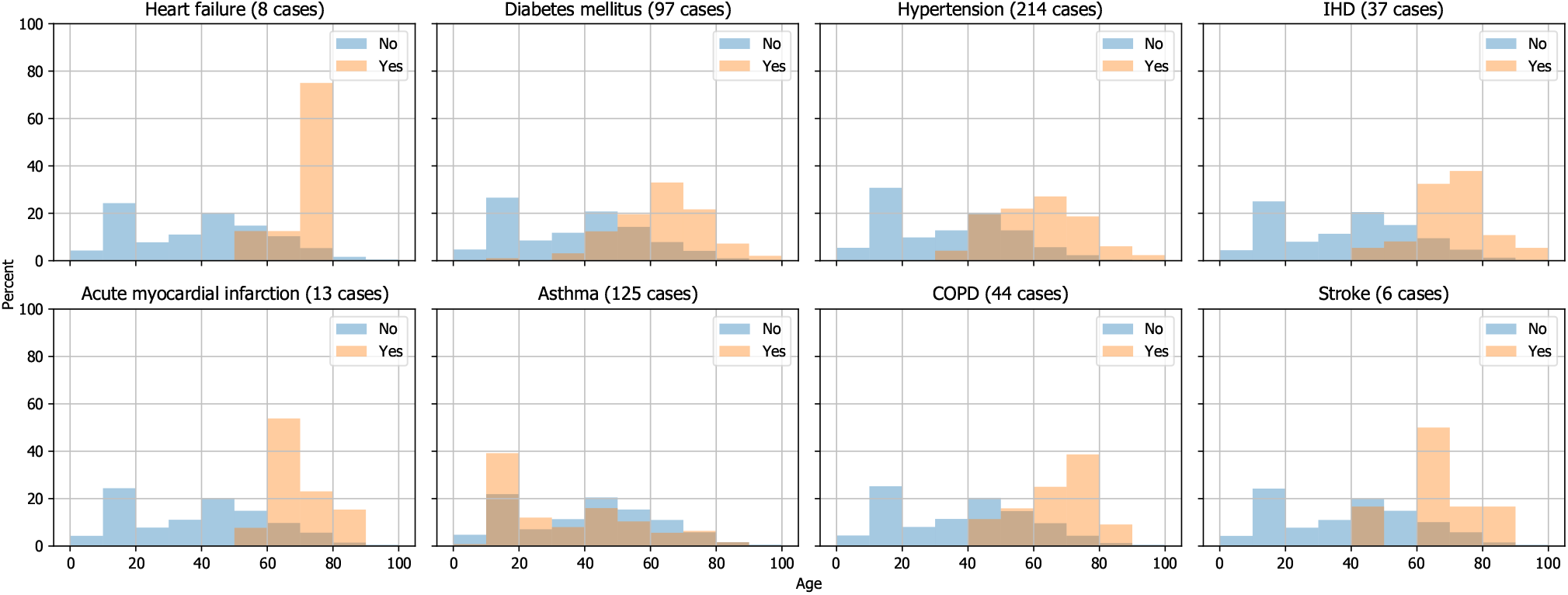
Density of patient comorbidities by age. Yes = at risk; No = not at risk.

**Figure 3.**
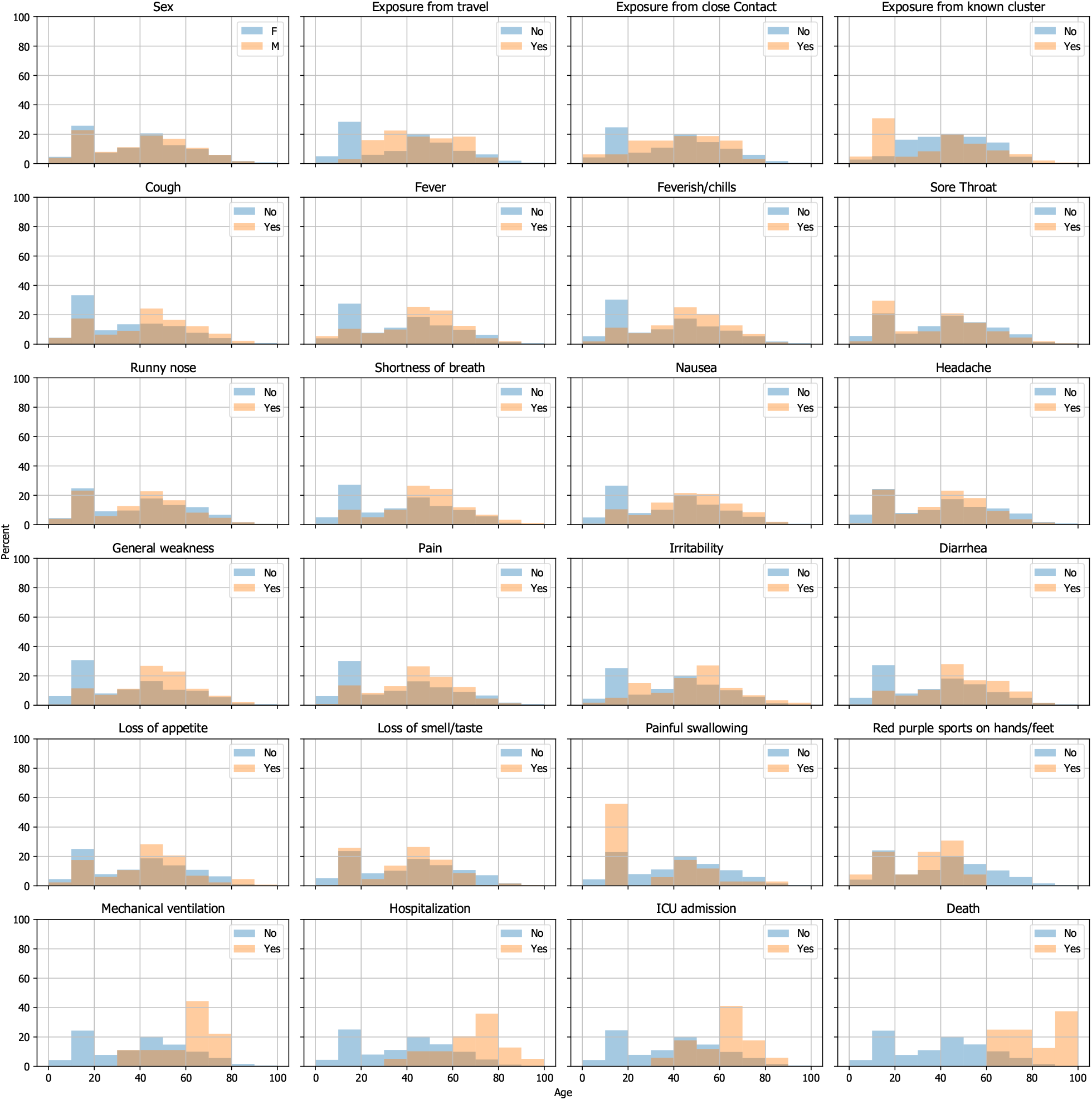
Density of patient sex, mode of infection acquisition, symptoms, and outcomes by age. Yes = at risk; No = not at risk.

## 4 Methods

We employ imbalance-aware supervised classification (AdaBoost and XGBoost) and unsupervised anomaly detection (OCSVM) algorithms to identify patients likely to experience severe COVID outcomes. Unsupervised anomaly detection is employed to detect both at-risk and deceased cases. However, the supervised models only detect the at-risk cases due to the severe class imbalance and too few records of deceased cases in the available dataset.

To gain statistical evidence regarding the models’ performance stability, we implement pipelines of five times repeated 5-fold cross-validation (CV) for tuning and evaluating each model. Slightly different versions of the repeated CV are implemented for supervised and unsupervised experiments settings. Both versions are discussed in detail during the following subsections. We additionally calculate SHAP values across all models to add model explainability to our framework.

### 4.1 Imbalance-aware supervised classification

We implement imbalance-aware AdaBoost and XGBoost models for binary classification of patients as either normal or at-risk. The models are provided with sample instances of both outcomes during the training. To manage the class imbalance in the data, both models take advantage of custom sample weights. For AdaBoost, we tune the penalty for mislabeling the positive (at-risk) samples to 15 times more than mislabeling negative samples, while for XGBoost we tune the penalty to 300 times more.

To go beyond simple CV and address possible performance stochasticity due to the small dataset size, we repeat 5-fold CV five times with different seeds for the randomized data splitting. Each experiment consists of 25 training and evaluation iterations. The performances from all these iterations are aggregated to obtain the optimal hyperparameters and validate the results. In each iteration of the supervised settings, we use stratified splitting to train and evaluate the models on 80% and 20% portions of the entire dataset, respectively.

### 4.2 Unsupervised anomaly detection

We implement two unsupervised anomaly detection OCSVM models to detect the at-risk cases and deceased cases as anomalies. The models only access the normal (not at-risk, not deceased) class during the training and learn to model the distribution of the normal population. After training, the models are able to classify samples from both the normal and at-risk (or deceased) populations as either normal or out-of-distribution (anomaly). Due to requiring samples only from the majority class during the training, OCSVM is well-suited for classification with severely imbalanced data, as in our COVID dataset.

Due to the small number of samples from the anomaly populations (42 (4%) at-risk, 8 (< 1%) deceased), we slightly modified the five times repeated 5-fold CV implementation. We isolate all anomaly samples at each iteration and split the data that only includes the majority samples into 80% and 20% train and test normal-only sets. The normal-only train set is used to train the models, and the normal-only test set is shuffled with all the anomaly samples to create the final test set. Then, similar to the supervised experiments, the results from the 25 iterations are aggregated for tuning and validation.

### 4.3 Interpretability

To integrate SHAP into the five times repeated 5-fold CV, we calculate a set of SHAP values for the test set samples during each iteration. To be able to later compare results between different models, at the end of each CV we normalize SHAP values across all samples and all features to have mean 0 and standard deviation 1. By the end of the 25 experiments, we have calculated five different sets of SHAP values per sample in the dataset. The results are aggregated for each sample by averaging the values across the five sets. We designate this set of average SHAP values as the final SHAP value set for the samples. In collaboration with our clinical partners, we also define five feature groups of clinical interest: constitutional symptoms, respiratory symptoms, fever symptoms, comorbidities group, and symptoms group (Table 1). Note that symptomatic feature is an individual feature from the training data, which indicates to the model whether or not a patient had experienced symptoms. On the other hand, the symptoms feature group is the bundle of all symptom-related features that allows us to study the correlated contribution of all members of the group. We calculate the sum of SHAP values for the features belonging to each group as the group’s marginal contribution to the output of the models.

## 5 Results

The mean and standard deviation of the true positive rate (TPR), false positive rate (FPR), F1 score [13], and AUC for each prediction tool are reported in Table 2 (see Appendix A for ROC curves; note that OC-SVM is not compatible with ROC curves). For AdaBoost and XGBoost, different discrimination thresholds yield different TPR/FPR results. For XGBoost, we derived the optimal discrimination thresholds by minimizing TPR*-* FPR. For the AdaBoost model, the default discrimination function implemented in the Python `scikit-learn` library [24] was optimal.

**Table 2:**
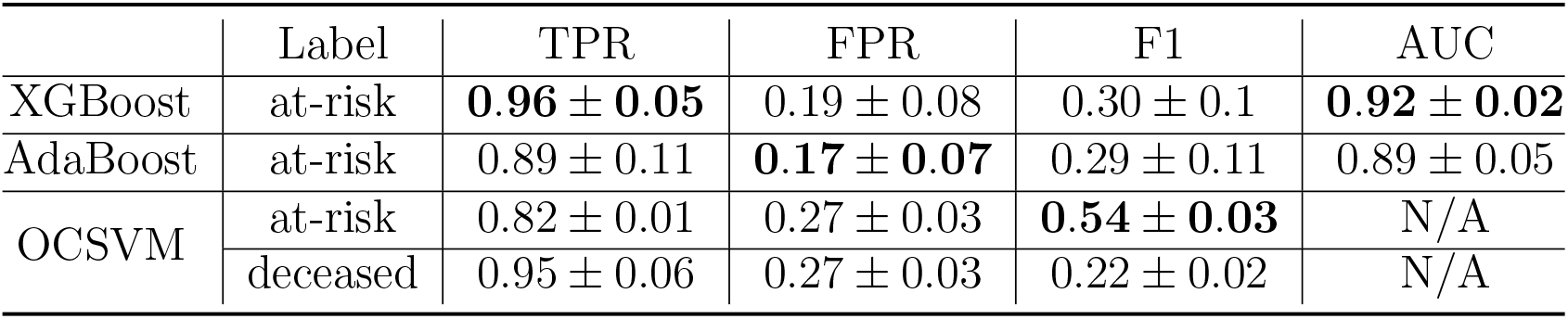
Performance metrics. Bold: best performance for at-risk prediction

|  | Label | TPR | FPR | F1 | AUC |
| --- | --- | --- | --- | --- | --- |
| XGBoost | at-risk | **$0.96 \pm 0.05$** | $0.19 \pm 0.08$ | $0.30 \pm 0.1$ | **$0.92 \pm 0.02$** |
| AdaBoost | at-risk | $0.89 \pm 0.11$ | **$0.17 \pm 0.07$** | $0.29 \pm 0.11$ | $0.89 \pm 0.05$ |
| OCSVM | at-risk | $0.82 \pm 0.01$ | $0.27 \pm 0.03$ | **$0.54 \pm 0.03$** | N/A |
| | deceased | $0.95 \pm 0.06$ | $0.27 \pm 0.03$ | $0.22 \pm 0.02$ | N/A |

Figures 4 and 5 illustrate the SHAP value features importances of each feature and feature group for at-risk and deceased prediction, respectively. The SHAP importances for XGBoost are shown here as it was the best performing predictor, however, the feature importances from the other models are generally in agreement with these importances (Appendix B), and we specifically observe that feature group importances for all methods are similar, relative to each other (Figure 6). Notably, symptoms as a group are consistently more important than comorbidities as a group. Furthermore, shortness of breath is the most important individual indicator for at-risk prediction, while being symptomatic is the most important individual indicator for death; in both predictions, the symptoms feature group is most important indicator overall. While age is an important indicator in both predictions, the relationship between high/low age and outcome is not always consistent, as evidenced by the mixed red/blue spots in the SHAP graphs.

**Figure 4.**
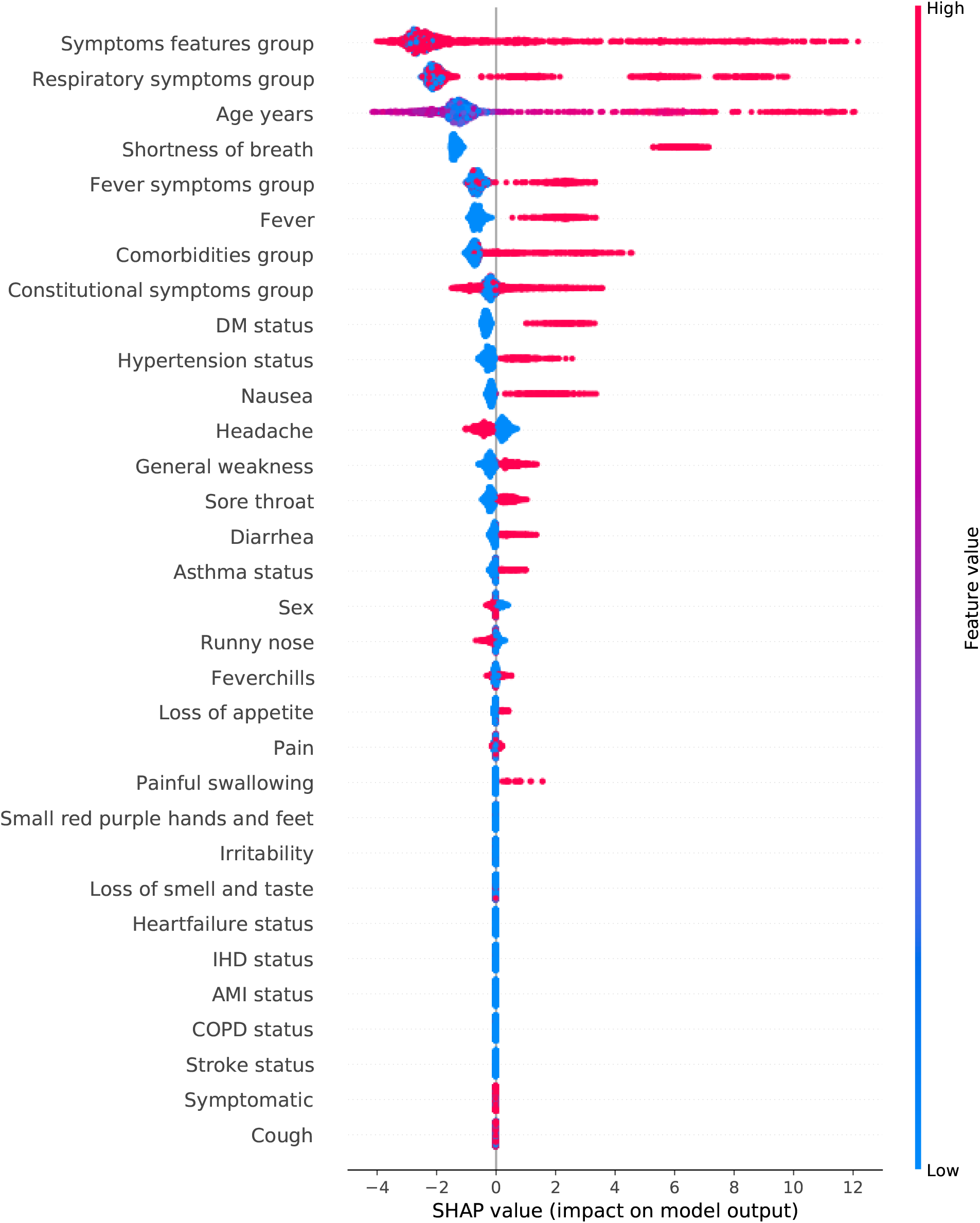
Scaled SHAP feature importance for XGBoost at-risk prediction

**Figure 5.**
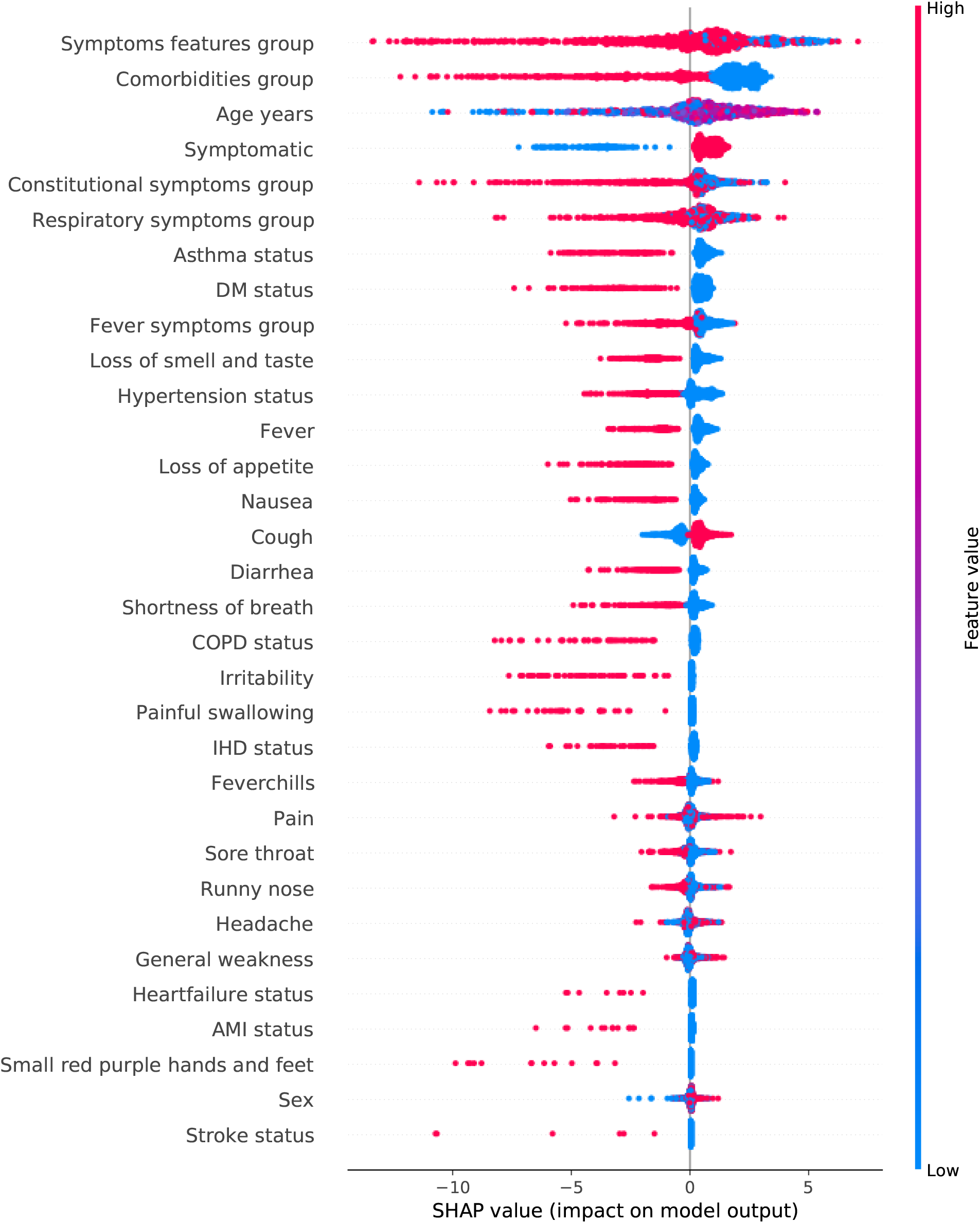
Scaled SHAP feature importance for OCSVM deceased prediction

**Figure 6.**
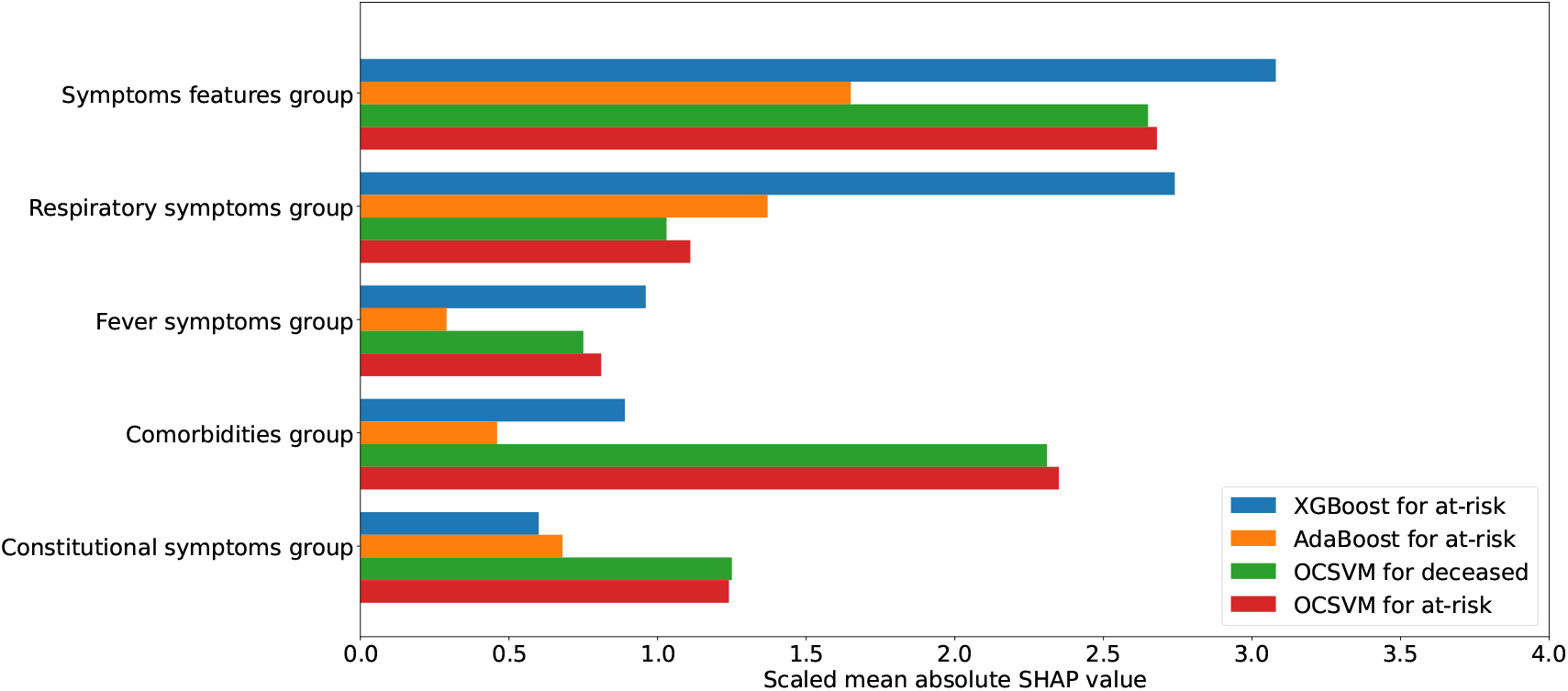
SHAP feature group importance for all predictors, after scaling SHAP values for all features to [0,1] within each predictor for visual clarity

These graphs also help validate the models’ understanding of what information is essential in the early prognosis of COVID patients. For example, in agreement with the common wisdom regarding COVID, the models identified age, shortness of breath, fever, and diabetes as the most significant individual risk indicators. Note that contrary to the AdaBoost and XGBoost models, higher feature values are associated with a negative impact on the model output for the OCSVM models due to the OCSVM implementation in `scikit-learn` which labels anomaly predictions as −1, while AdaBoost and XGBoost predict the at-risk labels as 1.

## 6 Discussion

The novelty of our study lies in combining transparency with significant predictive power despite minimal data, and is leveraged to make the first COVID patient outcome predictions at the time of RT-PCR testing. This work provides a generalizable early prognosis and rapid triaging framework that can be quickly adopted and integrated into the testing process to inform advanced resource allocation decisions in the event of a sudden and widespread disease outbreak similar to COVID.

Our approach does not depend on data that may be time-consuming or expensive to collect; in fact, our data can almost entirely self-reported by the patient, with the exception of fever status, which can be quickly collected during testing. Most machine learning literature on COVID prognosis takes advantage of accurate results from expensive diagnostic tools such as radiography imaging [20, 25, 30] or blood tests [1, 3, 29]. Such data is cumbersome to gather for those infected with SARS-CoV-2, and it may not be collected on a mass scale by default; further, this data can only be collected once the patient is in the hospital, at which point, advanced resource allocation is no longer possible. Instead, our framework puts little to no added stress on the system because it depends only on attributes such as age, minimal comorbidities background, and apparent symptoms at the time of diagnosis. Therefore, the framework is easily integratable into the current healthcare infrastructure at regional or national levels.

Further, we have shown significant predictive power, on par with the best performances among the literature, even with a small dataset of less than 1000 COVID patients. For at-risk prediction, we obtained an AUC of 92% with XGBoost, compared to 86% [29] and 92% [1], obtained with 18000 and 6995 patient records, respectively. Fewer studies train models specifically to predict mortality due to COVID. Subudhi et al. [29], Yan et al. [32] leverage feature-rich and large datasets to compare the performance of several supervised models for detecting COVID mortality from hospitalized patients and report high AUC measures (93% and 99%). Even though this performance is higher than our OCSVM, one should note that the performances are not fully comparable since our model is trained on a smaller dataset, for which only anomaly detection is an appropriate methodology, and predicts mortality at the time of the RT-PCR diagnosis, much earlier than hospitalization.

Additionally, we are using machine learning tools to go beyond mere predictions. On the micro-level, high performance and sample-level model explainability through SHAP provide clinical insight for each patient. On the macro-level, the aggregated sample-based explanations can guide assessment of risk factors in a local population, which provides further ability to allocate scarce resources appropriately.

The advantage of our framework lies in that the predictions are made at the time of RT-PCR positivity, and only a minimal number of such events are required for model training. In addition, because the framework is effective with only a small dataset, it can be applied to small regions to capture specific characteristics of the regional population and to accommodate changes regarding viral transmission factors and/or vaccination. However, it is important to note that the risk factors identified here are relevant only to the time period, population, and outcomes of the current study, and we again emphasize the lack of vaccination status data. For clinical implementation, a number of revisions must be taken into consideration, including collection of vaccination status, and possibly which vaccines were taken. Changes in the new virus variants or vaccination status may result in different constellations of symptoms and different degrees of severe infection rates. Therefore, the framework requires continuously updated data. Additionally, the occurrence of events such as hospitalization or ICU admission to some extent depends on the capacity and willingness of the local health care system to provide specialized care to patients.

## 7 Conclusion

We validated and explained the behavior of three machine learning models to predict, at the time of RT-PCR testing, whether COVID patients are at risk of severe outcome and/or death. This early prediction allows for advanced allocation of scarce healthcare resources, including ventilators and clinical staff. Our models were trained on minimal data from a small cohort of fewer than 1000 patients. The models are easy to use, can be adjusted based on regional needs, and showed significant and consistent predictive power in the prognosis of patients at risk. Finally, we incorporated our extended SHAP explainability into the framework to gain insights from the models’ behavior, allowing for clinical validation of their decisions. A future avenue of study is to validate the results and study the differences when applying the framework on multiple cohorts of geographically separated patients who may have significantly different underlying risks. Additionally, the models should be regularly updated with new cases, should contain vaccination status for improved accuracy, and their predictions should be compared with clinical predictions.

## Data Availability

Data is not available due to ethical requirements

## A ROC curves

**Figure 7.**
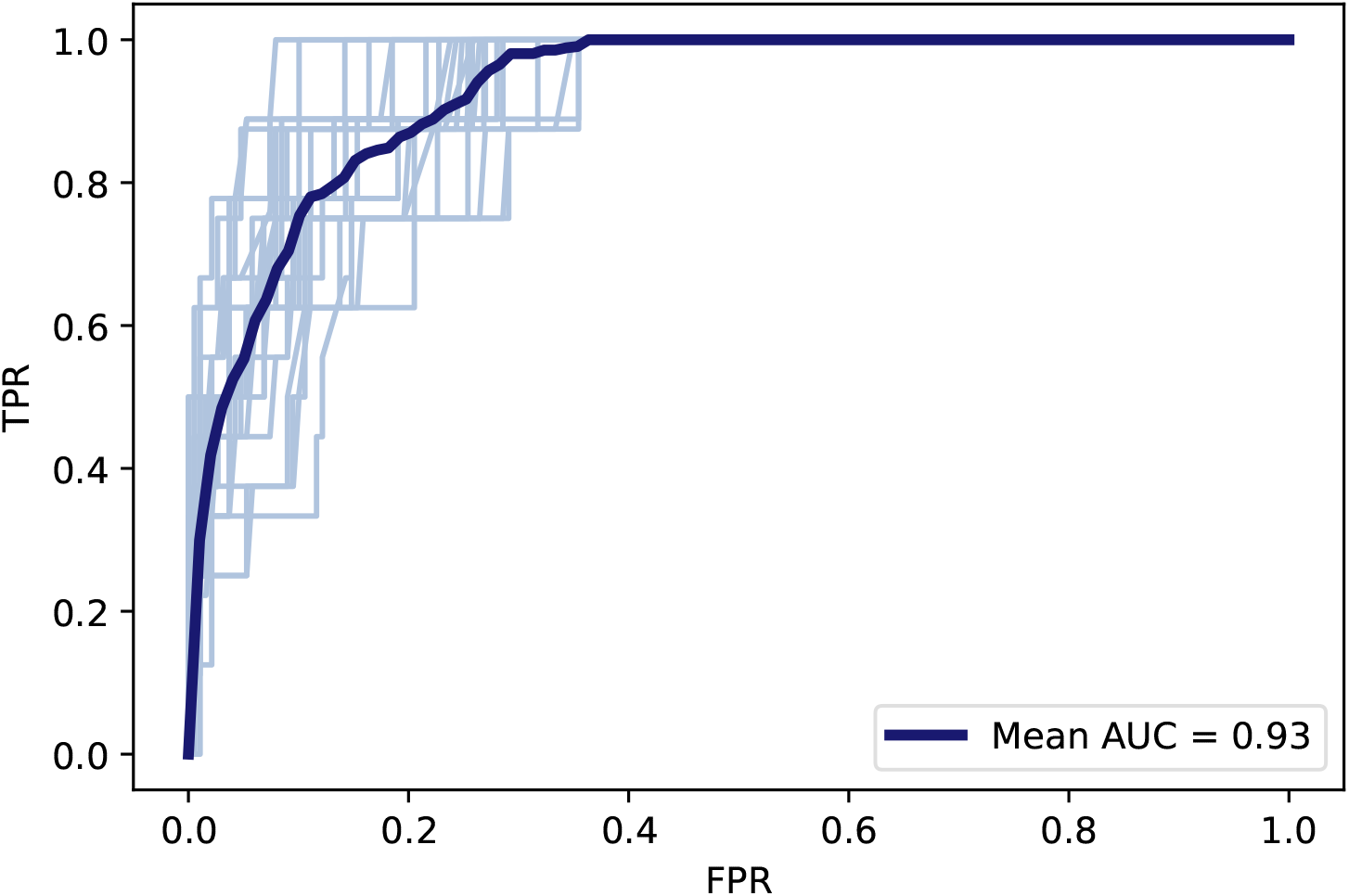
ROC curve for XGBoost at-risk prediction across all validation iterations; bold line is the average curve

**Figure 8.**
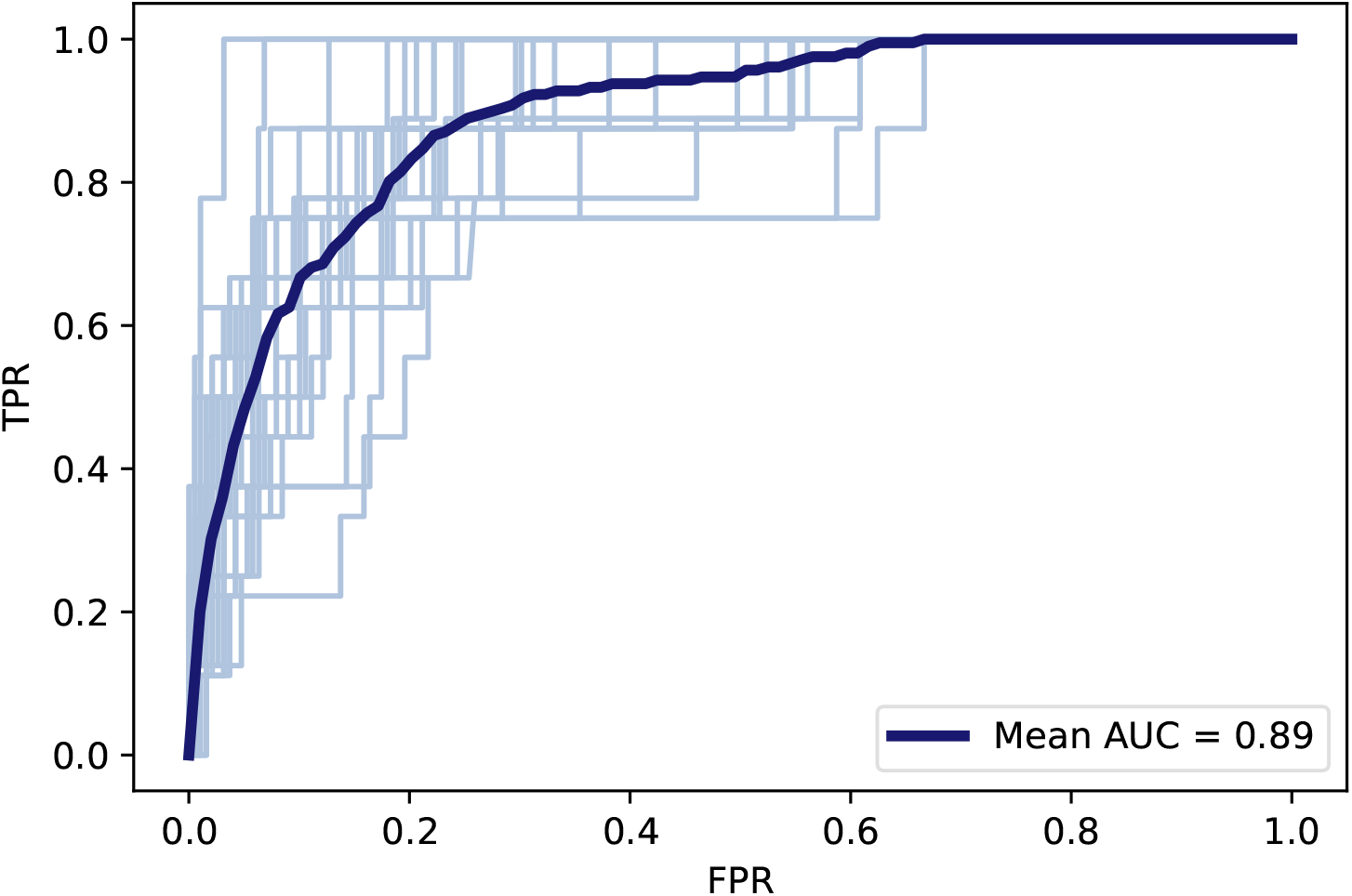
ROC curve for AdaBoost at-risk prediction across all validation iterations; bold line is the average curve

## B SHAP values for all methods

**Figure 9.**
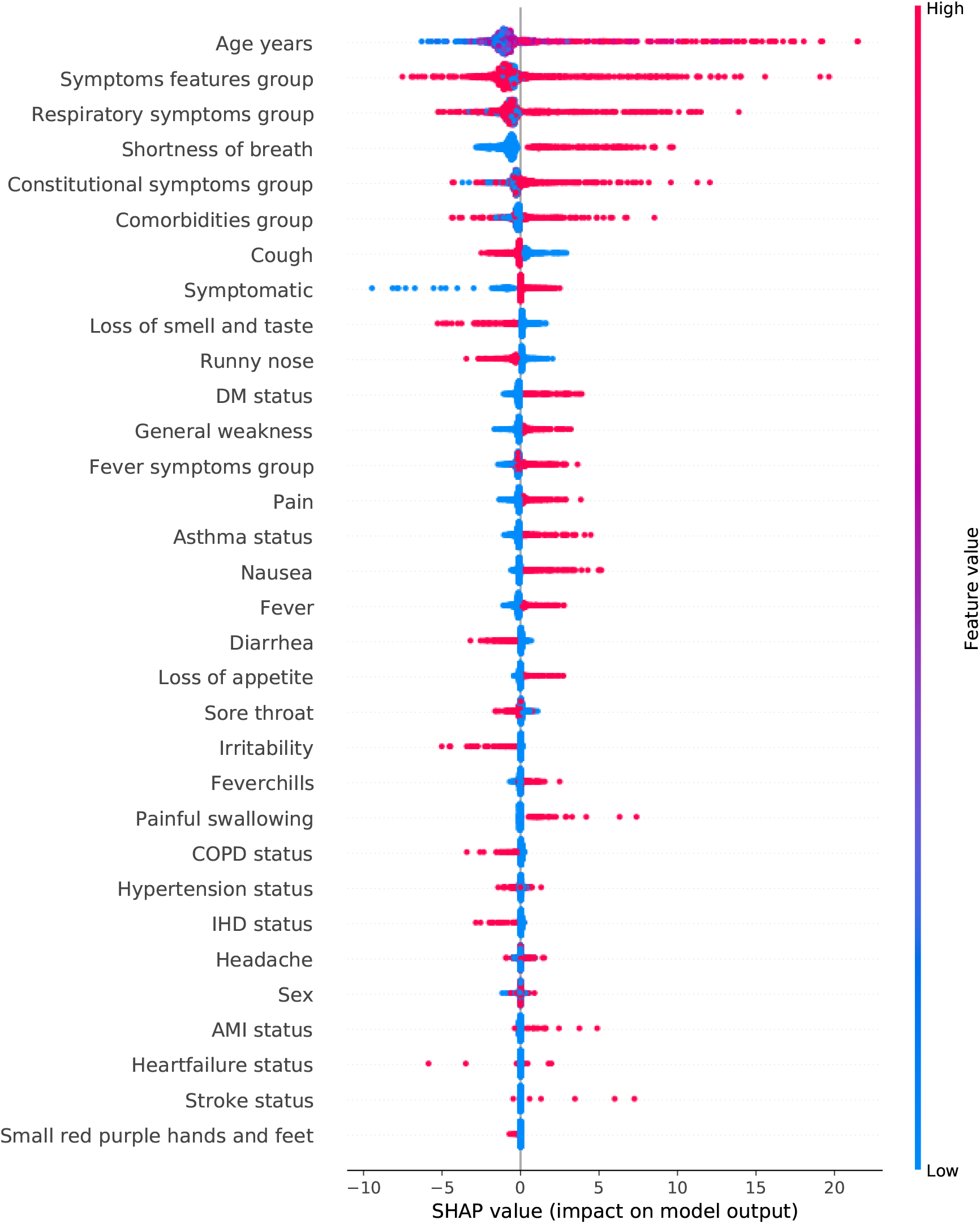
Scaled SHAP feature importance for AdaBoost at-risk prediction

**Figure 10.**
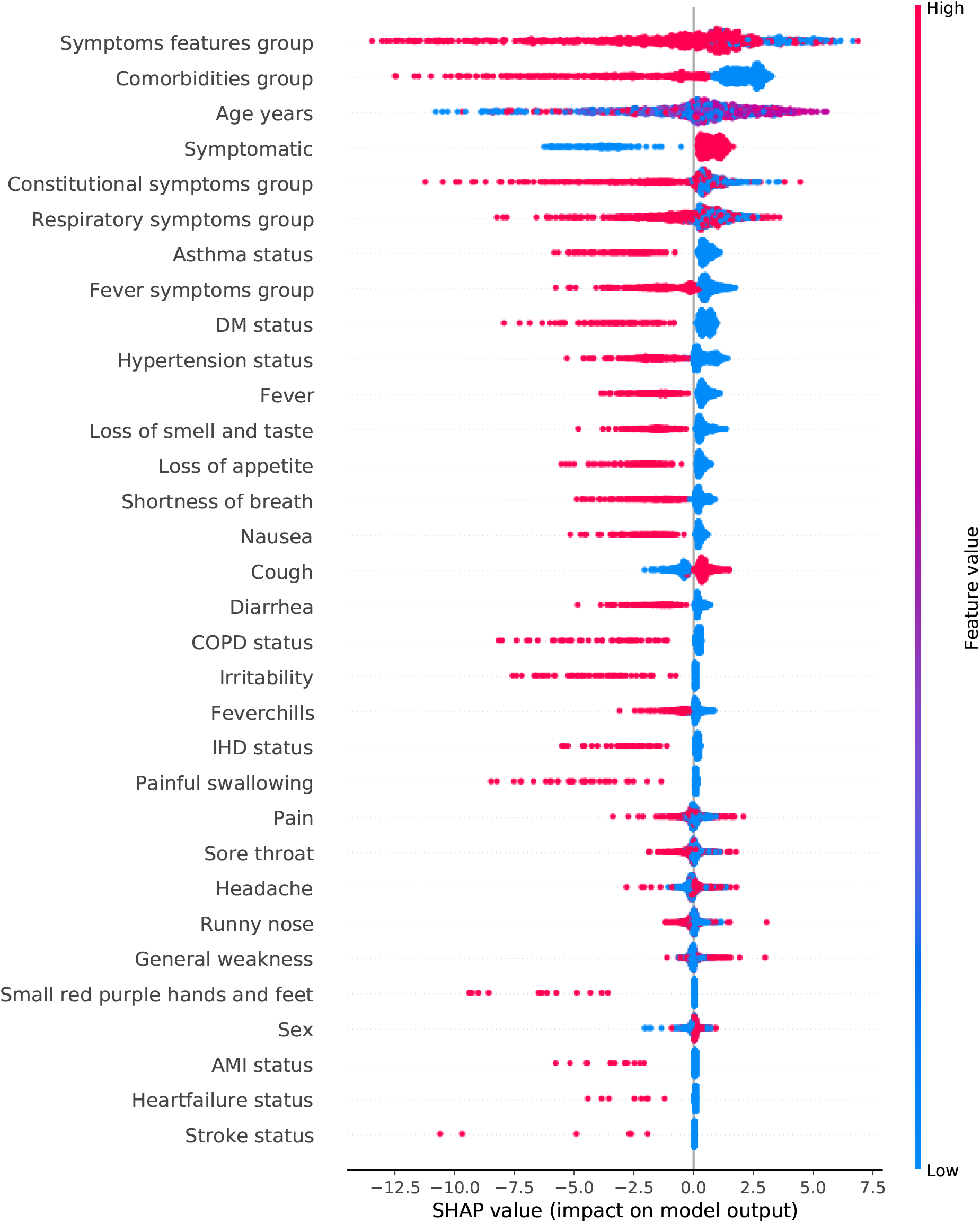
Scaled SHAP feature importance for OCSVM at-risk prediction

## C Optimized hyperparameter values

**Table 3:**
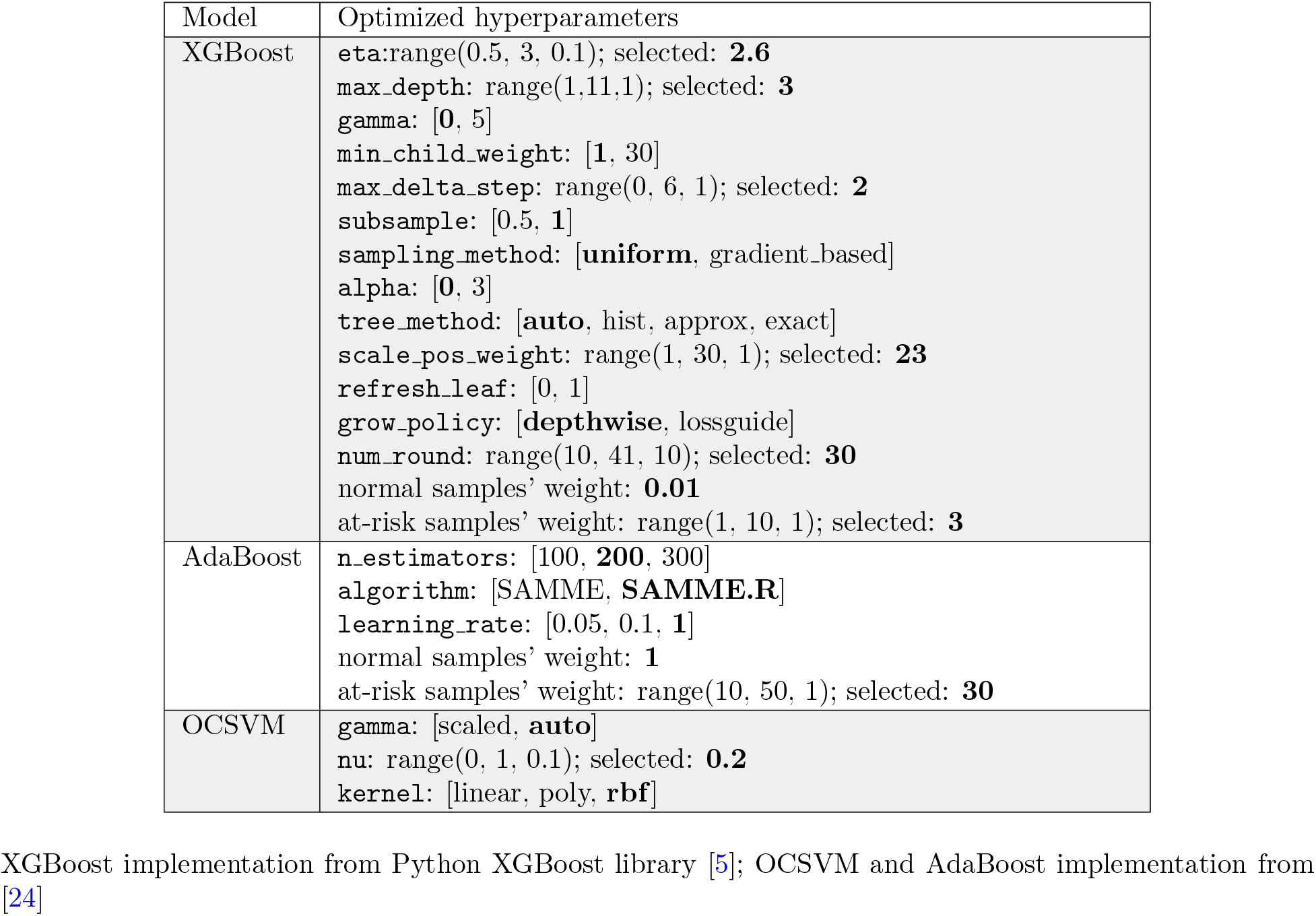
Optimized hyperparameters; bold is the optimal hyperparameter found.

